## Supplementary material for "Knowledge, attitude, and practice related to the COVID-19 pandemic among undergraduate medical students in Indonesia: a nationwide cross-sectional study"

**S1 Table.** Characteristics of the study population (n=4870)[a]

| Variable | N (%) | Knowledge | | Attitude | | Practice | |
|---|---|---|---|---|---|---|---|
| | | **Inadequate; 3418 (70.2)** | **Adequate; 1452 (29.8)** | **Inadequate; 1710 (35.1)** | **Adequate; 3160 (64.9)** | **Inadequate; 2360 (48.5)** | **Adequate; 2510 (51.5)** |
| Age (years) | 20 (19-21) | 20 (19-21) | 20 (19-22) | 20 (19-21) | 20 (19-21) | 20 (19-21) | 20 (19-21) |
| Sex | | | | | | | |
| Male | 1471 (30.2) | 1069 (72.7) | 402 (27.3) | 506 (34.4) | 965 (65.6) | 812 (55.2) | 659 (44.8) |
| Female | 3399 (69.8) | 2349 (69.1) | 1050 (30.9) | 1204 (35.4) | 2195 (64.6) | 1548 (45.5) | 1851 (54.5) |
| Location | N=4831 | | | | | | |
| Java | 4199 (86.9) | 2937 (69.9) | 1262 (30.1) | 1465 (34.9) | 2734 (65.1) | 2034 (48.4) | 2165 (51.6) |
| Sumatra | 237 (4.9) | 173 (73.0) | 64 (27.0) | 83 (35.0) | 154 (65.0) | 111 (46.8) | 126 (53.2) |
| Central Indonesia[b] | 103 (2.1) | 63 (61.2) | 40 (38.8) | 36 (35.0) | 67 (65.0) | 56 (54.4) | 47 (45.6) |
| Eastern Indonesia[c] | 292 (6.0) | 212 (72.6) | 80 (27.4) | 114 (39.0) | 178 (61.0) | 144 (49.3) | 148 (50.7) |
| Institution type | N=4831 | | | | | | |
| Public | 2525 (52.3) | 1705 (67.5) | 820 (32.5) | 859 (34.0) | 1666 (66.0) | 1325 (52.5) | 1200 (47.5) |
| Private | 2306 (47.7) | 1680 (72.9) | 626 (27.1) | 839 (36.4) | 1467 (63.6) | 1020 (44.2) | 1286 (55.8) |
| Academic level | | | | | | | |
| Pre-clinical | 3925 (80.6) | 2878 (73.3) | 1047 (26.7) | 1404 (35.8) | 2521 (64.2) | 1928 (49.1) | 1997 (50.9) |
| Clinical | 945 (19.4) | 540 (57.1) | 405 (42.9) | 306 (32.4) | 639 (67.6) | 432 (45.7) | 513 (54.3) |
| Living with[d] | | | | | | | |
| Family | 4154 (85.3) | 2930 (70.5) | 1224 (29.5) | 1448 (34.9) | 2706 (65.1) | 2020 (48.6) | 2134 (51.4) |
| Non-family | 58 (1.2) | 40 (69.0) | 18 (31.0) | 20 (34.5) | 38 (65.5) | 32 (55.2) | 26 (44.8) |
| Alone | 658 (13.5) | 448 (68.1) | 210 (31.9) | 242 (36.8) | 416 (63.2) | 308 (46.8) | 350 (53.2) |
| Number of housemate (people)[d] | 4 (3-5) | 4 (3-5) | 4 (3-5) | 4 (3-5) | 4 (3-5) | 4 (3-5) | 4 (3-5) |
| Living with children | | | | | | | |
| Yes | 2282 (46.9) | 1615 (70.8) | 667 (29.2) | 792 (34.7) | 1490 (65.3) | 1103 (48.3) | 1179 (51.7) |
| No | 2588 (53.1) | 1803 (69.7) | 785 (30.3) | 918 (35.5) | 1670 (64.5) | 1257 (48.6) | 1331 (51.4) |
| Living with elderly | | | | | | | |
| Yes | 1155 (23.7) | 786 (68.1) | 369 (31.9) | 394 (34.1) | 761 (65.9) | 563 (48.7) | 592 (51.3) |
| No | 3715 (76.3) | 2632 (70.8) | 1083 (29.2) | 1316 (35.4) | 2399 (64.6) | 1797 (48.4) | 1918 (51.6) |
| Marital status | | | | | | | |
| Married | 21 (0.4) | 15 (71.4) | 6 (28.6) | 6 (28.6) | 15 (71.4) | 7 (33.3) | 14 (66.7) |
| Not married | 4847 (99.5) | 3402 (70.2) | 1445 (29.8) | 1704 (35.2) | 3143 (64.8) | 2352 (48.5) | 2495 (51.5) |
| Divorced | 2 (0.0) | 1 (50.0) | 1 (50.0) | 0 (0.0) | 2 (100) | 1 (50.0) | 1 (50.0) |

| | | | | | | | |
|---|---|---|---|---|---|---|---|
| Family income | | | | | | | |
| ≤ IDR 1,500,000 | 241 (4.9) | 168 (69.7) | 73 (30.3) | 82 (34.0) | 159 (66.0) | 116 (48.1) | 125 (51.9) |
| IDR 1,500,001-2,500,000 | 273 (5.6) | 200 (73.3) | 73 (26.7) | 119 (43.6) | 154 (56.4) | 154 (56.4) | 119 (43.6) |
| IDR 2,500,001-3,500,000 | 534 (11.0) | 382 (71.5) | 152 (28.5) | 198 (37.1) | 336 (62.9) | 259 (48.5) | 275 (51.5) |
| > IDR 3,500,000 | 3822 (78.5) | 2668 (69.8) | 1154 (30.2) | 1311 (34.3) | 2511 (65.7) | 1831 (47.9) | 1991 (52.1) |
| History of chronic illness | | | | | | | |
| Yes | 316 (6.5) | 200 (63.3) | 116 (36.7) | 108 (34.2) | 208 (65.8) | 158 (50.0) | 159 (50.0) |
| No | 4554 (93.5) | 3218 (70.7)_ | 1336 (29.3) | 1602 (35.2) | 2952 (64.8) | 2202 (48.4) | 2352 (51.6) |
| Volunteered in health sectors | | | | | | | |
| Yes | 864 (17.7) | 533 (61.7) | 331 (38.3) | 258 (29.9) | 606 (70.1) | 373 (43.2) | 491 (56.8) |
| No | 4006 (82.3) | 2885 (72.0) | 1121 (28.0) | 1452 (36.2) | 2554 (63.8) | 1987 (49.6) | 2019 (50.4) |
| Volunteered in non-health sectors | | | | | | | |
| Yes | 3149 (64.7) | 2129 (67.6) | 1020 (32.4) | 1042 (33.1) | 2107 (66.9) | 1473 (46.8) | 1676 (53.2) |
| No | 1721 (35.3) | 1289 (74.9) | 432 (25.1) | 668 (38.8) | 1053 (61.2) | 887 (51.5) | 834 (48.5) |
| Family members diagnosed with COVID-19 | | | | | | | |
| Yes | 387 (7.9) | 258 (66.7) | 129 (33.3) | 99 (25.6) | 288 (74.4) | 177 (45.7) | 210 (54.3) |
| No | 3831 (78.7) | 2679 (69.9) | 1152 (30.1) | 1378 (36.0) | 2453 (64.0) | 1828 (47.7) | 2003 (52.3) |
| Don’t know | 652 (13.4) | 481 (73.8) | 171 (26.2) | 233 (35.7) | 419 (64.3) | 355 (54.4) | 297 (45.6) |
| Contacts with COVID-19 patients | | | | | | | |
| Yes | 167 (3.4) | 97 (58.1) | 70 (41.9) | 42 (25.1) | 125 (74.9) | 66 (39.5) | 101 (60.5) |
| No | 3618 (74.3) | 2532 (70.0) | 1086 (30.0) | 1289 (35.6) | 2329 (64.4) | 1694 (46.8) | 1924 (53.2) |
| Don’t know | 1085 (22.3) | 789 (72.7) | 296 (27.3) | 379 (34.9) | 706 (65.1) | 600 (55.3) | 485 (44.7) |
| Had been a COVID-19 patient | | | | | | | |
| Yes[e] | 33 (0.7) | 22 (66.7) | 11 (33.3) | 12 (36.4) | 21 (63.6) | 11 (33.3) | 22 (66.7) |
| No | 3990 (81.9) | 2784 (69.8) | 1206 (30.2) | 1423 (35.7) | 2567 (64.3) | 1869 (46.8) | 2121 (53.2) |
| Don’t know | 847 (17.4) | 612 (72.3) | 235 (27.7) | 275 (32.5) | 572 (67.5) | 480 (56.7) | 367 (43.3) |

[a]Unless explicitly stated, data are presented in n (%), mean ± standard deviation, or median (interquartile range). [b]Includes Sulawesi and Kalimantan. [c]Includes Bali, Nusa Tenggara, Maluku, and Papua. [d]Defined as the people living with the respondents at the time of questionnaire completion. [e]Includes both confirmed and unconfirmed (suspected or probable) cases. COVID-19, coronavirus disease 2019; IDR, Indonesian Rupiah

**S2 Table.** Item-specific responses on the participants' knowledge on COVID-19 (n=4870)

| Signaling question | Correct, n (%) |
|---|---|
| 1. Children and teenagers do not need to conduct preventive measures against COVID-19 | 4729 (97.1) |
| 2. SARS-CoV-2, the etiology of COVID-19, is transmitted via respiratory droplets of infected patients | 4697 (96.4) |
| 3. Which of the following can confirm the diagnosis of COVID-19? | 4631 (95.1) |
| 4. The main clinical findings in patients with COVID-19 are fever, fatigue, dry cough, and muscle tenderness (myalgia) | 4087 (83.9) |
| 5. Currently, not a single drug has been proven to cure COVID-19 | 4056 (83.3) |
| 6. In COVID-19 patients, what are the radiological findings that might be seen in the chest CT-scan result? | 3934 (80.8) |
| 7. Cold and nasal congestion are less likely to be found in patients with COVID-19 compared to patients with common cold | 2782 (57.1) |
| 8. What are the laboratory findings that may be present in COVID-19 patients? | 1548 (31.8) |
| 9. Eating or getting into contact with a wild animal might cause someone to get infected by the COVID-19 virus | 1548 (31.8) |
| 10. How long is the median incubation period of SARS-CoV-2 in human body? (in days) | 782 (16.1) |

COVID-19, coronavirus disease 2019; CT-scan, computed tomography scan.

**S3 Table.** Item-specific responses on the participants' attitude towards COVID-19 (n=4870)

| Signaling question | Strongly disagree | Disagree | Neutral | Agree | Strongly agree |
|---|---|---|---|---|---|
| 1. Face mask is effective to prevent COVID-19 infection | 46 (0.9) | 99 (2.0) | 391 (8.0) | 2389 (49.1) | 1945 (39.9) |
| 2. Washing your hands might prevent you from getting COVID-19 infection | 24 (0.5) | 15 (0.3) | 163 (3.3) | 2197 (45.1) | 2471 (50.7) |
| 3. Early detection of COVID-19 might improve the outcomes of treatment | 26 (0.5) | 33 (0.7) | 281 (5.8) | 2205 (45.3) | 2325 (47.7) |
| 4. It is possible to take care of COVID-19 patients in their home | 338 (6.9) | 762 (15.6) | 1636 (33.6) | 1655 (34.0) | 479 (9.8) |
| 5. Health education might reduce the rate of COVID-19 infection | 32 (0.7) | 38 (0.8) | 330 (6.8) | 2165 (44.5) | 2305 (47.3) |
| 6. COVID-19 is a serious disease | 27 (0.6) | 29 (0.6) | 305 (6.3) | 1888 (38.8) | 2621 (53.8) |
| 7. Once a vaccine for COVID-19 is available, I will voluntarily get vaccinated | 29 (0.6) | 42 (0.9) | 703 (14.4) | 1873 (38.5) | 2223 (45.6) |
| 8. COVID-19 is a curable disease | 39 (0.8) | 77 (1.6) | 808 (16.6) | 2362 (48.5) | 1584 (32.5) |
| 9. People living in my surroundings already have a good awareness regarding COVID-19 | 548 (11.3) | 1253 (25.7) | 1503 (30.9) | 1237 (25.4) | 329 (6.8) |
| 10. The government should have prohibited people from traveling to and from areas infected with COVID-19 to prevent transmission | 44 (0.9) | 89 (1.8) | 792 (16.3) | 1985 (40.8) | 1960 (40.2) |
| 11. If the number of COVID-19 cases continues to rise, the government should close public places/areas. (e.g., school, place of worship, shopping center, etc.) | 36 (0.7) | 70 (1.4) | 584 (12.0) | 1850 (38.0) | 2330 (47.8) |
| 12. If the number of COVID-19 cases continues to rise, the government should be ready to impose a lockdown policy | 51 (1.0) | 89 (1.8) | 644 (13.2) | 1726 (35.4) | 2360 (48.5) |

COVID-19, coronavirus disease 2019.

**S4 Table.** Item-specific responses on the participants' practice towards COVID-19 (n=4870)

| Signaling question | Strongly disagree | Disagree | Neutral | Agree | Strongly agree |
|---|---|---|---|---|---|
| 1. In this pandemic, I have always washed my hands for at least 20 seconds | 22 (0.5) | 63 (1.3) | 569 (11.7) | 2087 (42.9) | 2129 (43.7) |
| 2. In this pandemic, I have always tried to avoid touching my eyes, mouth, and mouth with my hand directly | 20 (0.4) | 181 (3.7) | 1163 (23.9) | 2176 (44.7) | 1330 (27.3) |
| 3. In this pandemic, I have always implemented a proper coughing and sneezing etiquette | 10 (0.2) | 24 (0.5) | 232 (4.8) | 1695 (34.8) | 2909 (59.7) |
| 4. In this pandemic, I have always tried to use disinfectants or alike-solutions to clean the surface of my surroundings | 41 (0.8) | 146 (3.0) | 594 (12.2) | 1802 (37.0) | 2287 (47.0) |
| 5. In this pandemic, I have always tried to use disinfectants or alike-solutions to clean my handphone | 146 (3.0) | 388 (8.0) | 1060 (21.8) | 1643 (33.7) | 1633 (33.5) |
| 6. In order to prevent contracting and spreading COVID-19, I avoid going out of my home | 57 (1.2) | 77 (1.6) | 474 (9.7) | 2179 (44.7) | 2083 (42.8) |
| 7. In this pandemic, I have always consumed vitamins to strengthen my immune system | 129 (2.6) | 279 (5.7) | 965 (19.8) | 1816 (37.3) | 1681 (34.5) |
| 8. In the last few days, I have consumed antibiotics or other non-herbal medications to prevent COVID-19 infection | 1927 (39.6) | 1118 (23.0) | 890 (18.3) | 525 (10.8) | 410 (8.4) |
| 9. In this pandemic, I have always put my face mask on before getting into contact with other people | 8 (0.2) | 14 (0.3) | 120 (2.5) | 1296 (26.6) | 3432 (70.5) |
| 10. In this pandemic, I have always avoided handshaking, hugging, or kissing | 17 (0.3) | 50 (1.0) | 322 (6.6) | 1754 (36.0) | 2727 (56.0) |
| 11. In this pandemic, I have always tried to avoid crowds and getting into close contact with people | 12 (0.2) | 20 (0.4) | 280 (5.7) | 2025 (41.6) | 2533 (52.0) |

| | | | | | |
|---|---|---|---|---|---|
| 12. In this pandemic, I have consumed herbal products or other traditional medicines (e.g., *jamu*, *temulawak*, and other herbal "drugs" intended to affect my health) | 1449 (29.8) | 990 (20.3) | 1056 (21.7) | 821 (16.9) | 554 (11.4) |

COVID-19, coronavirus disease 2019.

**S5 Table.** Correlation between the students' trust in health information sources with knowledge, attitude, and practice towards COVID-19 (n=4870)[a]

| Items | Knowledge | Attitude | Practice |
|---|---|---|---|
| Television | -0.001 | -0.005 | -0.007 |
| Newspaper | -0.005 | 0.003 | -0.010 |
| Online news | 0.008 | 0.007 | -0.026 |
| Social media | -0.009 | 0.000 | -0.040* |
| Government statement | 0.019 | -0.001 | 0.013 |
| Health institution statement | 0.012 | -0.004 | -0.002 |
| Expert opinion | 0.020 | -0.007 | -0.020 |

*p<0.01. [a]Analyzed with Spearman's correlation test. COVID-19, coronavirus disease 2019.
